# Feasibility of the Oldenburg phrase test (OLPHRA) in cochlear-implant users

**DOI:** 10.64898/2026.11.27.26368847

**Authors:** Annabell Urban, Inga Holube, Saskia Ibelings, Sebastian Puschmann, Andreas Radeloff, Pascale Sandmann

## Abstract

The Oldenburg Phrase Test (OLPHRA) is a novel speech-recognition test utilizing a fixed matrix structure of four-word everyday phrases delivered via a synthesized female voice. This prospective, observational study evaluated the feasibility and clinical utility of the OLPHRA during early rehabilitation and compared it against established clinical standards, namely the Freiburg Monosyllabic Speech Test (FMST) and the Oldenburg Sentence Test (OLSA). Fifteen newly implanted cochlear implant (CI) users were tracked longitudinally at one, two, and four weeks post-activation, and compared to a cohort of fourteen experienced CI users. To index cognitive demand and listening effort during active perception, verbal response times (VRT) and task-evoked pupil dilations were recorded simultaneously. Speech-recognition scores for the newly implanted cohort improved significantly over the four-week period, and did not differ significantly from those of the experienced users at week four, with ceiling effects for both matrix-style tests. However, FMST scores were significantly poorer in both groups. The OLPHRA evoked significantly shorter log-transformed VRTs than the OLSA, indicating faster behavioural processing. In the exploratory pupillometric analysis, a growth curve model revealed a significantly steeper initial pupillary slope for the OLPHRA than for the OLSA, which may indicate faster recruitment of cognitive resources during phrase encoding. Taken together, these findings support the clinical feasibility of the OLPHRA for tracking auditory rehabilitation in the early post-activation period. VRTs and pupil dilation showed diverging patterns that warrant further study. (trial registration: German Clinical Trials Register (DRKS), URL: https://www.drks.de/DRKS00040892, registration number: DRKS00040892)

## Introduction

A cochlear implant (CI) is a neural prosthesis that bypasses the damaged hair cells to electrically stimulate the remaining auditory nerve fibres [1–3], thereby improving speech recognition, social participation, and health-related quality of life [4]. In Germany, a structured multi-stage clinical guideline governs this interdisciplinary rehabilitation framework, combining speech processor programming, auditory training, and audiological evaluations [4,5]. This timeline comprises initial fitting, follow-up therapy, and long-term aftercare [5].

Audiometric speech-recognition tests serve as the primary metrics for evaluating this clinical progress. In Germany, the Freiburg Monosyllabic Speech Test (FMST) is a clinical standard in the early post-activation period. It consists of open-set monosyllabic words, thus offering little linguistic context [6]. In later rehabilitation stages, matrix tests, such as the Oldenburg Sentence Test (OLSA), are used in adaptive procedures that determine the speech-recognition threshold (SRT) in noise. The OLSA uses a fixed syntactic structure with semantically unpredictable sentences [7]. Recently, [8,9] developed the Oldenburg Phrase Test (OLPHRA) which retains the matrix layout of the OLSA but uses short everyday phrases embedded in a semantic context, spoken by a synthetic voice. The amount of context in speech material can be quantified by the j-factor [10]. It relates the probability of recognizing a whole speech item (sentence or phrase) to the probability of recognizing its individual word, and can be interpreted as the effective number of independent words per item. The stronger the contextual coupling between words, the lower the j-factor. The OLPHRA has a lower j-factor than the OLSA [9,11].

Speech-recognition tasks assess more than simple peripheral sensitivity. They also place explicit demands on working memory and cognitive capacities [12]. The Framework for Understanding Effortful Listening (FUEL) defines this allocation of mental resources as listening effort [12]. According to the “Ease of Language Understanding (ELU) Model”, optimal speech perception proceeds via rapid, automatic, implicit lexical matching with long-term memory representations [13]. However, when speech is degraded by noise or peripheral pathology, processing shifts to explicit, resource-demanding mechanisms that recruit working memory to reconstruct the message [13]. For CI users, this signal degradation is continuous. The CI-induced Resource Capture (CIRC) Model suggests that decoding restricted spectro-temporal cues requires sustained central resource allocation [14]. Consequently, CI users can exhibit elevated listening effort even when standard clinical speech-recognition scores appear satisfactory [15].

To quantify these cognitive demands, research applies behavioural and physiological metrics alongside subjective self-reports [16]. Verbal Response Time (VRT), measured within a single-task paradigm, indexes processing speed and cognitive load by capturing the temporal latency between stimulus offset and participants’ vocalisation onset alongside behavioural accuracy [17,18]. VRT is sensitive to listening difficulty. Lower signal-to-noise ratios (SNRs), fluctuating maskers, advanced age, and hearing impairment systematically prolong response latencies [19–21]. However, questions remain regarding the specific cognitive sub-dimensions VRT isolates and its exact alignment with subjective self-reports of effort scales. While [18] did not find a close link between VRT and subjective effort scales, [19] demonstrated significant correlations between the two measures.

Behavioural measures (such as VRT) and physiological indices offer complementary insights into listening effort. Pupillometry provides a time-resolved physiological index of autonomic nervous system arousal during speech processing [22–24]. Task-evoked pupil dilation reflects continuous resource allocation and is typically evaluated via baseline size, mean pupil dilation, peak amplitude, and peak pupil latency [25]. Task-evoked pupillary responses show a high test-retest reliability across variations in speech-recognition performance [24,26]. While traditional research focused on peak metrics, other investigations utilized the initial pupillary slope following the stimulus onset as a marker of the immediate auditory processing demand during signal encoding with a steeper slope suggesting higher demands [21,27,28]. Furthermore, stimulus architecture directly shapes these physiological measures. Meaningful sentence structures evoke distinct pupil dilation compared to semantically unpredictable matrix tests in CI users, suggesting that test design alters central retrieval strategies [29].

This study aimed to investigate the feasibility and clinical utility of the OLPHRA in CI recipients during the early post-activation period, together with VRTs and pupillometry. We compared speech recognition in the OLPHRA with that in the OLSA and the FMST representing different rehabilitation stages and different amounts of linguistic context. Two cohorts of CI recipients were recruited: newly implanted users (CI-rehab) and experienced users (CI-experienced). The CI-rehab group was measured at three time points (one, two, and four weeks post-activation), whereas the CI-experienced group was measured once. To control for the contribution of the non-test ear across heterogeneous device configurations, a fixed contralateral masking noise was applied. Because accuracy alone does not fully describe speech processing, we simultaneously recorded VRTs and task-evoked pupil dilation. Pupillometric data were analysed with Growth Curve Analysis (GCA), similar to [21]. Owing to substantial data loss, this analysis was exploratory. We hypothesized that:

a. The predictable semantic structure of the OLPHRA will yield higher speech-recognition scores, shorter processing latencies (VRTs), and decreased pupillary slope (less processing demand) compared to the unpredictable matrix structure of the OLSA.
b. Speech-recognition performance across all test materials will exhibit systematic improvements over the first four weeks of device use for the CI-rehab group.
c. The cognitive load, indexed by VRT and pupillary slopes, will decrease as a function of both longitudinal rehabilitation time and the contextual availability of the speech materials.

## Methods

### Participants

Two groups of CI users were recruited for this study: a CI rehabilitation group (CI-rehab; *n* = 15, age: mean (SD) 65.2 (13.3) years; 5 male, 10 female) who had recently received their implant, and a CI-experienced group (*n* = 14, age: mean (SD) 51.0 (15.2) years; 8 male, 6 female) with at least two years of CI experience. Both groups included individuals with bimodal (CI and contralateral hearing aid; CI-rehab: *n* = 11; CI-experienced: *n* = 3), bilateral (two CIs; CI-rehab: *n* = 3; CI-experienced: *n* = 8), or single-sided deafness (SSD; CI-rehab: *n* = 1; CI-experienced: *n* = 3) configurations. Age, gender, fitting, duration of hearing loss, CI experience, and implanted devices of all participants are listed in Table 1. All participants were native German speakers. Based on existing clinical records, we excluded individuals with cognitive impairments or attention deficits, or those taking medications known to affect the parasympathetic nervous system [25,26,30]. All participants were recruited and tested at one centre. The sample comprised all eligible patients (29 patients were eligible; all agreed; 15 + 14 were included) of the larger clinical projects, and no sample size calculation was performed. Participants were recruited between 01^st^ July 2024 and 30^th^ September 2025.

**Table 1:** Participant’s information about gender, age, tested ear, fitting, duration of hearing loss, CI-experience, and CI-type.

| ID | Gender | Group | Age range (years) | Ear | Fitting | Hearing loss (years) | CI Experience (years) | CI type |
|---|---|---|---|---|---|---|---|---|
| 01 | Male | Rehab | 71-75 | Left | Bimodal | 5 | – | MED-EL, Sonnet 2 |
| 02 | Female | Rehab | 76-80 | Right | Bimodal | > 40 | – | Advanced Bionics, HiRes Ultra 3D |
| 03 | Female | Rehab | 46-50 | Left | Bimodal | < 1 | – | MED-EL, Sonnet 2 |
| 04 | Male | Rehab | 76-80 | Right | Bimodal | 11 | – | Cochlear, Kanso 2 |
| 05 | Male | Rehab | 56-60 | Right | Bimodal | 17 | – | Cochlear, CP1110 |
| 06 | Female | Rehab | 66-70 | Right | Bimodal | 14 | – | MED-EL, Sonnet 2 |
| 07 | Female | Rehab | 71-75 | Left | Bimodal | 9 | – | MED-EL, Sonnet 2 |
| 08 | Female | Rehab | 36-40 | Right | Bimodal | 6 | – | Cochlear, CP1110 |
| 09 | Male | Rehab | 56-60 | Left | SSD | 18 | – | MED-EL, Sonnet 2 |
| 10 | Female | Rehab | 71-75 | Right | Bilateral | 35 | – | Cochlear, CP1110 |
| 11 | Female | Rehab | 46-50 | Right | Bilateral | > 40 | – | Cochlear, Kanso 3 |
| 12 | Female | Rehab | 56-60 | Right | Bimodal | 10 | – | Cochlear, Kanso 3 |
| 13 | Female | Rehab | 76-80 | Left | Bilateral | 35 | – | Cochlear, CP1110 |
| 14 | Male | Rehab | 66-70 | Right | Bimodal | > 40 | – | MED-EL, Sonnet 3 |
| 15 | Female | Rehab | 76-80 | Right | Bimodal | 14 | – | Cochlear, CP1100 |
| 16 | Male | Experienced | 56-60 | Left | SSD | 10 | 8 | Cochlear, CP1100 |
| 17 | Male | Experienced | 21-25 | Right | Bilateral | 21 | 21 | Cochlear, CP1100 |
| 18 | Male | Experienced | 51-55 | Right | SSD | 6 | 5 | Advanced Bionics, HiRes Ultra 3D |
| 19 | Male | Experienced | 61-65 | Left | Bilateral | 34 | 8 | Cochlear, CP1000 |
| 20 | Female | Experienced | 66-70 | Right | Bilateral | 15 | 8 | Cochlear, CP1000 |
| 21 | Female | Experienced | 71-75 | Left | Bilateral | – | 10 | MED-EL, Sonnet 2 |
| 22 | Female | Experienced | 51-55 | Left | Bilateral | 16 | 8 | Cochlear, CP1000 |
| 23 | Male | Experienced | 56-60 | Left | Bimodal | 21 | 9 | Cochlear, CP1000 |
| 24 | Female | Experienced | 56-60 | Right | Bilateral | > 40 | 18 | Cochlear, CP1000 |
| 25 | Male | Experienced | 56-60 | Left | Bilateral | > 40 | 7 | Cochlear, CP1100 |
| 26 | Male | Experienced | 16-20 | Left | Bimodal | 17 | 8 | Cochlear, CP1100 |
| 27 | Male | Experienced | 51-55 | Left | SSD | 3 | 2 | MED-EL, Sonnet 2 |
| 28 | Female | Experienced | 46-50 | Right | Bilateral | 26 | 4 | MED-EL, Sonnet 2 |
| 29 | Female | Experienced | 31-35 | Right | Bimodal | 21 | 12 | Cochlear, N8 |

Abbreviations: CI, cochlear implant; SSD, single-sided deafness; –, not applicable or not available.

The present study extends a larger clinical project comparing the FMST and OLSA in the early and late post-activation period (not published yet). The CI-rehab participants were recruited within that project, and the CI-experienced cohort from the clinical routine. Because the FMST and OLSA were measured with the same setup in both projects, these data also contributed to the larger project. The OLPHRA, VRT recordings, and pupillometry were added for the present study. The study protocol was approved by the Medical Ethics Committee of Carl von Ossietzky University Oldenburg (approval number: 2021-095). The study was conducted in accordance with the Declaration of Helsinki, and all participants gave written informed consent. The study was registered retrospectively in the German Clinical Trials Register (DRKS00040892). Registration was completed after data collection had started, because the OLPHRA, VRT and pupillometry measurements were added as an extension to an existing clinical project that had not been registered as a trial.

### Speech-Recognition Tests

To evaluate speech recognition across varying degrees of contextual and lexical complexity, this study used three different speech audiometry materials: FMST, OLSA, and OLPHRA. The FMST consists of 20 lists of 20 open-set monosyllabic words spoken by a natural male voice to assess suprathreshold speech discrimination [6,31,32]. The OLSA utilizes a fixed, five-word syntactical matrix layout (name-verb-numeral-adjective-noun) with a closed-set vocabulary that remains semantically unpredictable (“Nina painted eleven white rings”; German: “Nina malt elf weiße Ringe”; [7,11,33]). Its sentences are spoken by a natural male speaker. The test also provides a corresponding speech-shaped masking noise (OLSAnoise) which is generated by a randomised temporal overlay of the speech material [7,11,33]. The OLPHRA consists of four-word everyday phrases with semantic context (article-adjective-noun-infinitive) generated with a synthesized female voice (“to watch the little cat”; German: “die kleine Katze beobachten”). Like the OLSA, it includes a corresponding speech-shaped masking noise (OLPHRAnoise; [8,9]). The j-factor of the OLSA is 4.29 at -5 dB SNR [7,8], compared to a maximum of 5 for a five-word sentence. For the OLPHRA, the j-factor is 3.1 at -6 dB SNR, compared to a maximum of 4 for a four-word phrase [8].

Percent-correct hit rates were converted to rationalized arcsine units (RAU) [34] using MATLAB 2022b (MathWorks, Natick, Massachusetts, USA) to analyse speech-recognition performance on a linear scale. This yielded speech-recognition scores in RAU for each speech test, group, and time point.

### Experimental Setup

Speech testing took place at the end of a routine clinical appointment. The CI-rehab group was tested following their standard 3-hour rehabilitation session (including audiometric testing, auditory training, and device fitting), whereas the CI-experienced group was tested after their annual 2-hour audiological assessment. The FMST was administered by qualified clinical personnel during the standard clinical routine using an Auritec AT1000 audiometer routed to an Auritec loudspeaker (Auritec, Hamburg, Germany). One test list was presented at 65 dB SPL in quiet in a sound-treated booth, with the loudspeaker at an azimuth of ±45° on the side of the CI and an ear-to-loudspeaker distance of 73 cm. The loudspeaker was calibrated for the participant’s position.

The OLSA and OLPHRA were tested inside a separate sound-treated booth. The Genelec 8030A loudspeaker (Genelec, Iisalmi, Finland) was also positioned at an azimuth of ±45° on the side of the CI, at an ear-to-loudspeaker distance of 64 cm. As for the FMST, the loudspeaker was calibrated for the participant’s position. Stimuli were presented at a fixed level of 65 dB SPL via the research versions of both speech tests in MATLAB 2017a. Fig 1 (left panel) shows a schematic of this setup. Concurrently, a separate laptop running Audacity (Muse Group, Limassol, Cyprus; version 2.4.2) presented a continuous masking noise (OLSAnoise or OLPHRAnoise, depending on the test) contralaterally at 75 dB SPL via Sennheiser HDA 300 circumaural headphones (Sennheiser, Wedemark, Germany). The headphones were calibrated prior to data collection.

**Fig 1:**
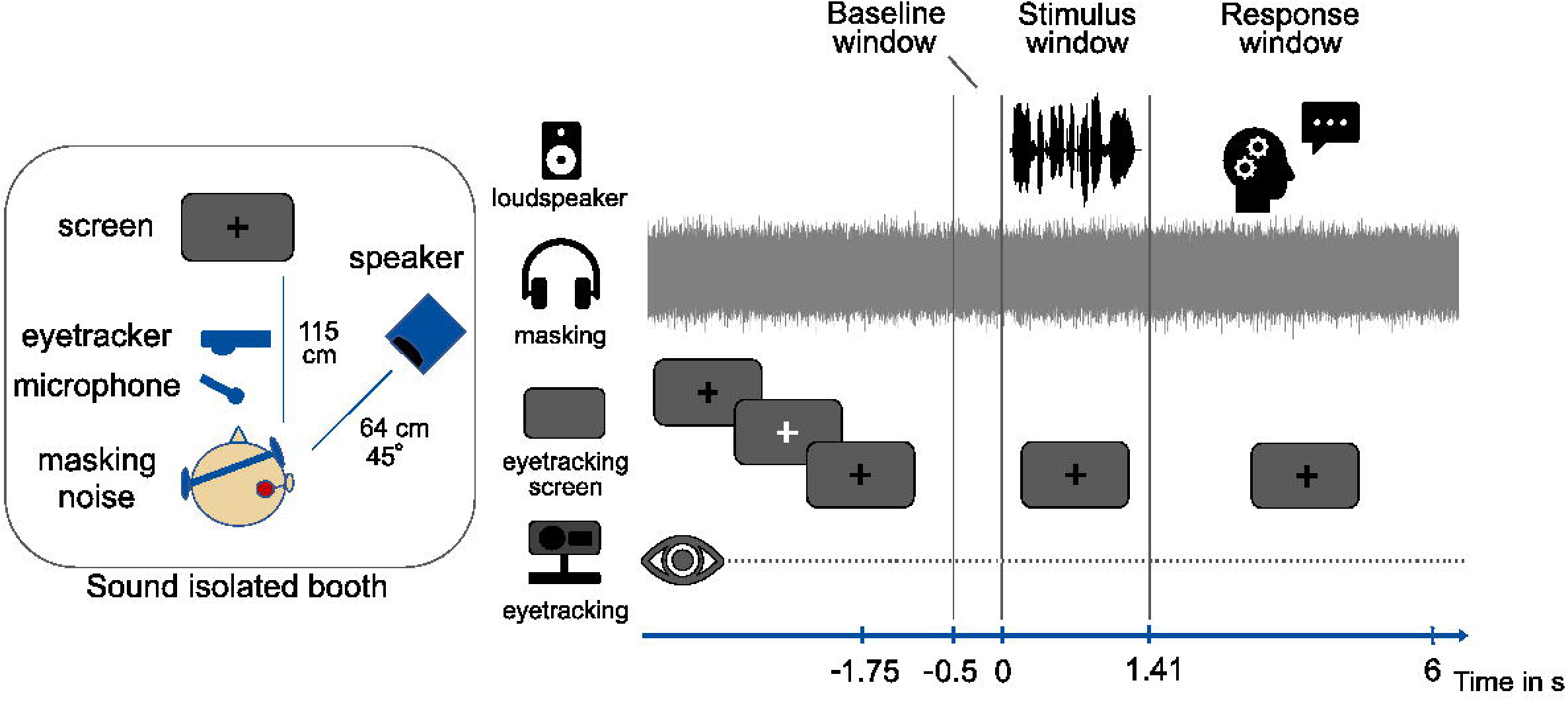
Experimental setup and trial sequence. Left: Schematic visualisation of the distances and positions of all devices inside the booth. Right: General measurement procedure within each trial.

### Procedure

The CI-rehab group was measured at three time points (one, two, and four weeks post-activation), whereas the CI-experienced group, serving as a reference cohort, performed the measurement protocol once. To minimize order effects, the order of OLSA and OLPHRA was randomized across participants. At each time point and for each test (OLPHRA and OLSA) participants completed two training lists: one in the best-aided condition (bimodal: with hearing aid; bilateral: with second CI, SSD: with normal-hearing ear) and one with the contralateral masking noise without contralateral device. These were followed by one measurement list per test in the latter condition (contralateral masking, no contralateral device). Training and measurement lists were randomly selected from the available lists of the respective test. The contralateral masking noise was set to 75 dB SPL to ensure that participants with SSD, low and moderate hearing loss could not hear the sentences or phrases from the loudspeaker with their normal-hearing ear.

The sequence of events within a single trial is shown in Fig 1 (right panel). As in [21], no post-stimulus retention interval was included, allowing for the simultaneous capture of transient pupillary responses and VRTs. Participants were instructed to repeat the sentence or phrase immediately after stimulus offset. The next stimulus started 2–3 s after the participant’s vocalisation, so as not to interfere with the normal course of a speech test procedure.

At the end of each measurement list, participants performed a two-alternative forced-choice (2AFC-) task on whether the speech material was synthetic or natural. They also rated their perceived listening effort for that test and time point on a scale from 1 (“no effort”; German: “gar nicht anstrengend”) to 5 (“extreme effort”; German: “äußerst anstrengend”).

### Verbal Response Times (VRT)

To capture behavioural processing speed alongside speech-recognition accuracy, verbal responses were recorded using a Shure SM58 microphone (Shure, Niles, Illinois, USA) positioned directly in front of the participant inside the booth. The sampling rates (*f*_s_) were set to the original rates of the speech material (OLSA: 32 kHz, OLPHRA: 44.1 kHz). VRTs were defined as the time elapsed between stimulus offset and onset of the participant’s vocalisation. Vocalisation onset was detected automatically with MATLABs detectSpeech() function.

VRT data preprocessing was executed in MATLAB 2022b. Mean VRTs were calculated for each measurement list and timepoint. To ensure that the timing metrics reflected successful lexical access rather than listening errors or confusion, mean VRTs were calculated using only the trials where at least one word was correctly repeated. If no word was repeated correctly in the measurement list, no VRT could be computed for that list, and the value was treated as missing. Following the methodology of [19], VRT data were log-transformed using the natural logarithm ln, with VRT expressed in ms. No minimum VRT reference threshold was applied because no vocalisation onset occurred during active stimulus presentation. This procedure provided a measure of log-transformed mean VRTs per test and per time point.

### Pupillometry

To track continuous cognitive workload during speech perception, pupillometric data were recorded inside the sound-treated booth under regulated ambient lighting conditions (mean illuminance: 30.54 lux; [25,35]). Participants sat centrally facing a visual display at a viewing distance of approximately 115 cm (see Fig 1, left panel). Eye-tracker control from outside the booth was implemented in MATLAB 2017a using Psychtoolbox [36]. Pupil size was recorded continuously from the right eye at *f*_s_ = 1000 Hz with the EyeLink 1000 Plus system (SR Research, Ottawa, Ontario, Canada).

Prior to each test list, participants performed a 9-point eye-tracking calibration. Each trial began with a grey background screen (RGB: 90, 90, 90) featuring a central black fixation cross. At 1.75 s prior to auditory stimulus onset, the fixation cross flashed white for 48 ms to signal the start of a new trial (see Fig 1, right panel).

Raw pupil data were processed via the pupil_pipeline toolbox [37] in MATLAB 2022b following standard guidelines [25]. Blinks and saccades were identified and linearly interpolated from 50 ms before to 150 ms after the event. The interpolated traces were filtered using a 10 Hz low-pass filter and epoched into 6-s windows. We applied a subtractive baseline correction based on a 500 ms window preceding stimulus onset, and EyeLink recordings in arbitrary units were converted to pupil size in mm with an artificial pupil calibration. Individual trials were averaged per participant, time point and test. Trials with more than 25% data loss due to artifact rejection were discarded, and participants with fewer than 50% valid trials remaining were excluded from further analysis [25].

Additionally, during the initial phase of data collection, a high-contrast visual cue caused systematic, stimulus-independent pupil elevation and was therefore removed from the experimental protocol. Consequently, the datasets affected by the systematic error and too few valid trials were excluded from the analysis. Of the 15 CI-rehab participants, 13 had valid pupil data at the four-week time point, and only 6 had valid data at all three time points (9/15, 60%, excluded from the longitudinal analysis). Of the 14 CI-experienced participants, 10 had valid data (4/14, 29%, excluded). Because the longitudinal sample (*n* = 6) was too small, the CI-rehab group was analysed at week four only. The final sample for the pupillometric analysis therefore comprised 23 participants (13 CI-rehab at four weeks, 10 CI-experienced). The analysis is therefore exploratory.

To isolate listening demand during active perception and separate it from motor responses (which can contaminate the signal and complicate interpretation), the pupillometric analysis was restricted to the stimulus presentation window. Consequently, the duration of the presented sentences was taken into account when defining the analysis window. The minimum, maximum, mean, and standard deviation of the sentence and phrase lengths are detailed in Table 2. We defined a conservative analysis window spanning 0 to 1.41 s, corresponding to the shortest overall stimulus duration recorded across speech materials. This physiological processing pipeline yielded the mean pupil dilation time series and its corresponding initial pupillary slope within the defined stimulus window.

**Table 2:** Minimum and maximum length of sentences from the OLSA and phrases from the OLPHRA, as well as mean and standard deviation (SD).

|  | **Oldenburg Sentence Test<br>(OLSA)** | **Oldenburg Phrase Test<br>(OLPHRA)** |
|---|---|---|
| minimum length in s | 1.77 | 1.41 |
| maximum length in s | 2.77 | 2.91 |
| Mean (SD) in s | 2.18 (0.21) | 1.96 (0.26) |

### Statistical Analyses

All statistical analyses were performed in R (version 4.5.2; R Core Team) using RStudio (Posit PBC, Boston, Massachusetts, USA) with two-sided tests and a significance level of α =.05. Descriptive statistics are given as mean and standard deviation (SD) with t-based 95% confidence intervals, or as median and interquartile range (IQR) where stated. Speech-recognition scores are illustrated in a growth-curve line plot, VRTs are presented as individual values and group means with 95% confidence intervals. Normality was assessed with the Shapiro-Wilk test. For the linear mixed models, homoscedasticity and normality of the residuals were checked visually with residual-versus-fitted plots and quantile-quantile plots. Outliers were identified using and were not excluded from the analyses using the 1.5 × IQR criterion (rstatix). Age differences between the groups were tested with a Mann-Whitney U test, because the age distribution of the CI-experienced group deviated from normality (Shapiro-Wilk: CI-experienced *p* = .030). Package versions: rstatix 0.7.3, lme4 1.1-38, lmerTest 3.2-0, emmeans 2.0.4. The preprocessed data and the analysis scripts (for statistical analysis and plots) are available at Zenodo and 10.5281/zenodo.23727483.

Differences in speech recognition were evaluated using repeated-measures analyses of variance (rm-ANOVAs) with the *rstatix* package. For within-group changes in the CI-rehab cohort, the rm-ANOVA models included time point (one, two, vs. four weeks) and test type (OLPHRA, OLSA, FMST) as within-subject factors. Between-group variations were examined via mixed ANOVAs using the CI-rehab four-week data, with group as the between-subject factor and test type as the within-subject factor. Sphericity was assessed with Mauchly’s test, and Greenhouse-Geisser corrections were applied where necessary. Effect sizes are reported as generalized eta squared (η²G).

Due to poor speech recognition, VRTs could not be calculated for 15 out of the 90 CI-rehab measurements and therefore were analysed with linear mixed-effects models (*lme4*, [38]; *lmerTest*, [39]) using all available values. Natural-log-transformed VRTs (ln of VRT in ms) were modelled with time point, test (OLPHRA, OLSA), and their interaction as fixed effects and a random intercept per participant. Degrees of freedom and *p* values for fixed effects were obtained with Satterthwaite’s approximation [39,40], which accounts for the uncertainty in the estimated variance components and has been shown to control the Type I error rate in mixed models [41]. Estimated differences are reported as ratios (back-transformed from the log scale) with 95% confidence intervals, obtained with the *emmeans* package [42]. To compare VRTs between groups, a similar model was fitted to the ln-transformed VRTs of the four-week CI-rehab measurement and the CI-experienced group. Group, test, and their interaction were entered as fixed effects with a random intercept of each participant. Participants without a valid VRT in a test were excluded (Tables 5 and 6 in the Results section). Contrasts and ratios were calculated as in the CI-rehab model. Full model outputs are given in S1 Table.

Post hoc pairwise comparisons (paired t-tests for the rm-ANOVAs, estimated marginal mean contrasts for the mixed models) were corrected for multiple comparisons with the Benjamini-Hochberg procedure, including the test contrasts across the three time points; 95% confidence intervals are unadjusted. The overall test contrast is a single comparison and was not adjusted. The association between the objective measure VRT and the subjective ratings of listening effort was assessed with Spearman’s rank correlation. Benjamini-Hochberg correction was applied within each group across the VRT correlations (6 tests in CI-rehab, 2 in CI-experienced).

Pupillometric data were analysed in an exploratory manner. Within the 0-1.41 s window, pupil traces were averaged into 100 ms bins and modelled with a GCA [43] using linear mixed-effects models (*lme4*, [38]; *lmerTest*, [39]), as in previous studies [21,25]. Based on visual inspection of the pupil traces, the time course was modelled as a first-order polynomial (linear trend). The primary model included time, test (OLPHRA, OLSA), group (CI-rehab at four weeks, CI-experienced), and all their interactions as fixed effects, plus age (grand-mean centred) and its interaction with time. Age was included because the groups differed significantly in age. The CI-rehab group was significantly older than the CI-experienced group (median [IQR]: 69 [57–75.5] vs. 55 [50.5–59] years; Mann-Whitney *U* = 158.5, *p* = .021). Test and group were sum-coded. Random effects were an intercept and a time slope for each participant × test combination, and an additional time slope for each participant. This structure fitted equivalently to a model with an additional random participant intercept (χ²(2) = 1.37, *p* = .50) but without a singular fit. *p* values were based on Satterthwaite degrees of freedom; model-estimated slopes and 95% confidence intervals were obtained with Kenward-Roger approximation [44] in the emmeans package [42]. In a sensitivity analysis, the model was refitted without age terms. The full model outputs are given in S2 Table. Additionally, mean pupil dilation and slope (linear fit) were calculated per participant and test as descriptive measures (Tables 7 and 8). As a secondary analysis, mean pupil dilation was analysed with a linear mixed model including test, group, their interaction, and age as fixed effects and a random intercept per participant. The association between mean pupil dilation and slope was assessed with Pearson’s correlation. The output of the mean-dilation model is given in S3 Table.

## Results

### Speech recognition

Descriptive statistics (mean (SD) with 95% confidence intervals) of hit rate and RAU are given in Table 3 for the CI-rehab group, and in Table 4 for the CI-experienced group.

**Table 3:** Speech-recognition performance of the CI-rehab group for each test (FMST, OLPHRA, OLSA) and time point (one, two, four weeks after initial fitting). Hit rates in % and RAU are given as mean (SD) and 95% confidence interval across participants (*n* = 15).

| **Time point** | **Test** | **Mean (SD) hit rate (%)** | **Confidence Interval (%)** | **Mean (SD) hit rate (RAU)** | **Confidence Interval (RAU)** |
|---|---|---|---|---|---|
| 1 | FMST | 18.7 (24.9) | [4.9, 32.5] | 13.5 (29.2) | [-2.7, 29.7] |
| 1 | OLPHRA | 42.3 (36.9) | [21.9, 62.8] | 37.6 (46.6) | [12.0, 63.1] |
| 1 | OLSA | 41.6 (33.8) | [22.9, 60.3] | 39.7 (40.7) | [17.2, 62.3] |
| 2 | FMST | 28.0 (25.3) | [14.0, 42.0] | 24.0 (30.2) | [7.2, 40.7] |
| 2 | OLPHRA | 58.9 (38.1) | [37.8, 80.0] | 57.7 (46.7) | [31.9, 83.6] |
| 2 | OLSA | 63.1 (34.1) | [44.3, 82.0] | 63.0 (41.5) | [40.0, 86.0] |
| 4 | FMST | 49.3 (30.6) | [32.4, 66.3] | 47.7 (33.2) | [29.3, 66.1] |
| 4 | OLPHRA | 77.1 (33.5) | [58.5, 95.6] | 79.3 (42.4) | [55.8, 102.8] |
| 4 | OLSA | 76.3 (27.4) | [61.1, 91.5] | 78.8 (34.5) | [59.7, 98.0] |

**Table 4:** Speech-recognition performance of the CI-experienced group for each test (FMST, OLPHRA, OLSA). Hit rates in % and RAU are given as mean (SD) and 95% confidence interval across participants (*n* = 14).

| **Test** | **Mean (SD) hit rate (%)** | **Confidence Interval (%)** | **Mean (SD) hit rate (RAU)** | **Confidence Interval (RAU)** |
|---|---|---|---|---|
| FMST | 67.9 (19.4) | [56.7, 79.1] | 67.7 (19.3) | [56.4, 78.7] |
| OLPHRA | 89.7 (15.9) | [80.5, 98.9] | 96.9 (20.4) | [85.1, 108.7] |
| OLSA | 87.8 (13.0) | [80.3, 95.3] | 94.0 (19.4) | [82.8, 105.2] |

The CI-rehab group demonstrated significant longitudinal improvements in speech-recognition performance over the first month of device use (Fig 2, left panel). The rm-ANOVA revealed significant main effects for both time point [*F*(2,28) = 35.7, *p* < .001, η²G = 0.15] and test [*F*(2,28) = 24.6, *p* < .001, η²G = 0.13]. The interaction between time point and test was not significant [*F*(2.6,36.3) = 0.99, *p* = .40, η²G = 0.004; Greenhouse-Geisser corrected], indicating that performance improved at a comparable rate across all three speech tests. Post-hoc tests confirmed that scores improved significantly from week one to week two (*p* < .001) and further increased from week two to week four (*p* < .001). Across all time points, scores for the FMST were significantly lower than those obtained for both the OLPHRA (*p* < .001) and OLSA (*p* < .001), whereas OLPHRA and OLSA scores did not differ significantly (*p* = .47).

**Fig 2:**
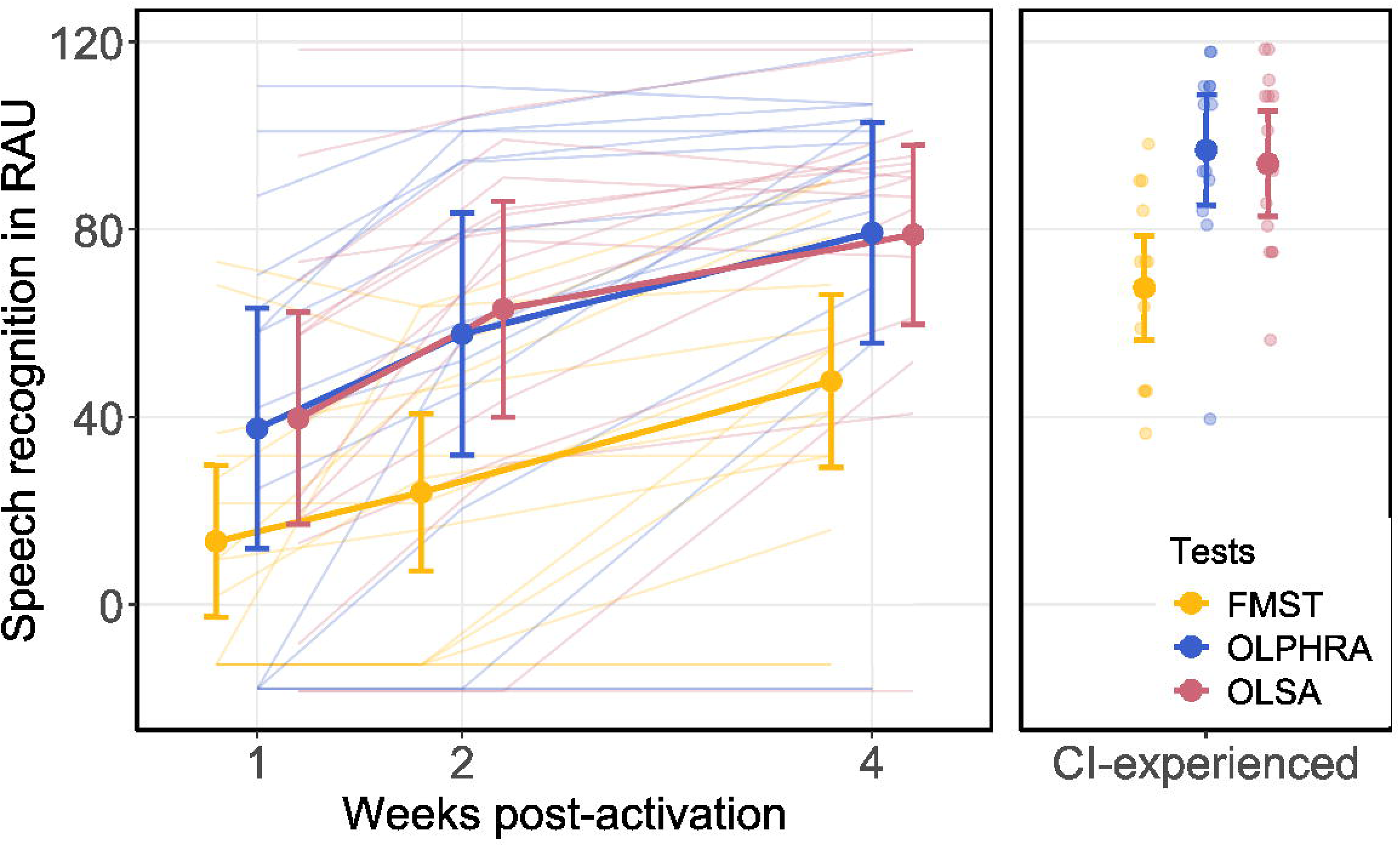
Speech-recognition scores (mean, confidence intervals) in RAU for the CI-rehab group (n=15; left panel) across different time points (one, two, four weeks after initial fitting) and for the CI-experienced group (n=14; right panel) at one timepoint. Statistical results and significant post-hoc comparisons after Benjamini-Hochberg correction were omitted for clarity and are reported in the Results section.

To evaluate performance relative to long-term users, we compared the four-week data of the CI-rehab group against the CI-experienced cohort (Fig 2, right panel). This between-group analysis revealed no significant interaction between group and test [*F*(2,54) = 0.15, *p* = .86, η²G = 0.001], and no significant main effect of group [*F*(1,27) = 3.11, *p* = .09, η²G = 0.08]. The main effect of test remained significant [*F*(2,54) = 31.7, *p* < .001, η²G = 0.19]. As in the within-group analysis, post-hoc tests showed significantly poorer performance for the FMST than for the OLPHRA (*p* < .001) and OLSA (*p* < .001), and no differences between OLSA and OLPHRA (*p* = .69). Scores for both matrix tests appear to be near maximum accuracy (RAU = 118.4) for the CI-experienced cohort (Fig 2, right panel), indicating a ceiling effect. Subjective categorization ratings regarding the voice of the matrix tests revealed a general ambiguity across both cohorts. In the CI-rehab group at the four-week time point, 8/15 (53.3%) participants correctly categorized the OLPHRA as synthesized speech, while 8/15 (53.3%) correctly rated the OLSA as natural. In the CI-experienced group, 8/14 (57.1%) correctly identified the OLPHRA as synthetic, whereas a higher proportion of 10/14 (71.4%) correctly categorized the OLSA as a natural voice.

### Verbal Response Time

Descriptive statistics (mean (SD) with 95% CI) of the VRTs are given in Table 5 for the CI-rehab group and in Table 6 for the CI-experienced group, both in seconds and as natural-log-transformed values (ln of VRT in ms; see Methods). VRTs could not be computed for 15 of 90 CI-rehab measurements because no word was repeated correctly. These were mostly at week one (see Table 5 for the number of valid datasets per condition). The mixed model therefore included 75 observations from 14 participants.

**Table 5:** VRTs of the CI-rehab group for each test (OLPHRA, OLSA) and time point (one, two, four weeks after initial fitting). VRTs in s and after logarithmic transformation in ln(ms) are given as mean (SD) and 95% confidence interval across valid datasets.

| **Time point** | **Valid datasets** | **Test** | **Mean (SD) VRT (s)** | **Confidence Interval (s)** | **Mean (SD) VRT (ln(ms))** | **Confidence Interval (ln(ms))** |
|---|---|---|---|---|---|---|
| 1 | n = 10 | OLPHRA | 1.1 (0.4) | [0.9, 1.3] | 6.9 (0.3) | [6.7, 7.1] |
| 1 | n = 13 | OLSA | 1.3 (0.7) | [1.0, 1.7] | 7.1 (0.4) | [6.9, 7.3] |
| 2 | n = 12 | OLPHRA | 1.0 (0.5) | [0.7, 1.3] | 6.8 (0.4) | [6.6, 7.1] |
| 2 | n = 13 | OLSA | 1.2 (0.4) | [1.0, 1.4] | 7.0 (0.4) | [6.8, 7.2] |
| 4 | n = 13 | OLPHRA | 0.8 (0.2) | [0.7, 0.9] | 6.6 (0.3) | [6.5, 6.8] |
| 4 | n = 14 | OLSA | 1.0 (0.3) | [0.9, 1.2] | 6.9 (0.3) | [6.8, 7.1] |

In the CI-rehab group, VRTs decreased significantly over time [*F*(2, 57.1) = 8.14, *p* < .001] and were shorter for the OLPHRA than for the OLSA [*F*(1, 57.3) = 10.30, *p* = .002; Fig 3, left panel]. Averaged across time points, OLPHRA VRTs were about 17% shorter than OLSA VRTs (ratio of geometric mean VRTs = 0.83, 95% confidence interval [0.74, 0.93]). The interaction between time point and test was not significant [*F*(2, 56.5) = 0.60, *p* = .55]. The direction of the difference was the same at all time points, but it was significant only at week four (ratio = 0.77, 95% confidence interval [0.64, 0.93], *p* = .023), not at week one (0.90, [0.73, 1.11], *p* = .303) or week two (0.83, [0.68, 1.01], *p* = .096). VRTs one week post activation were 1.32 times longer than four weeks post-activation (95% confidence interval [1.15, 1.52], *p* < .001), and VRTs two weeks post activation were 1.19 times longer than four weeks post-activation ([1.04, 1.37], *p* = .020). Weeks one and two did not differ significantly (ratio 1.11, [0.96, 1.28], *p* = .149).

**Fig 3:**
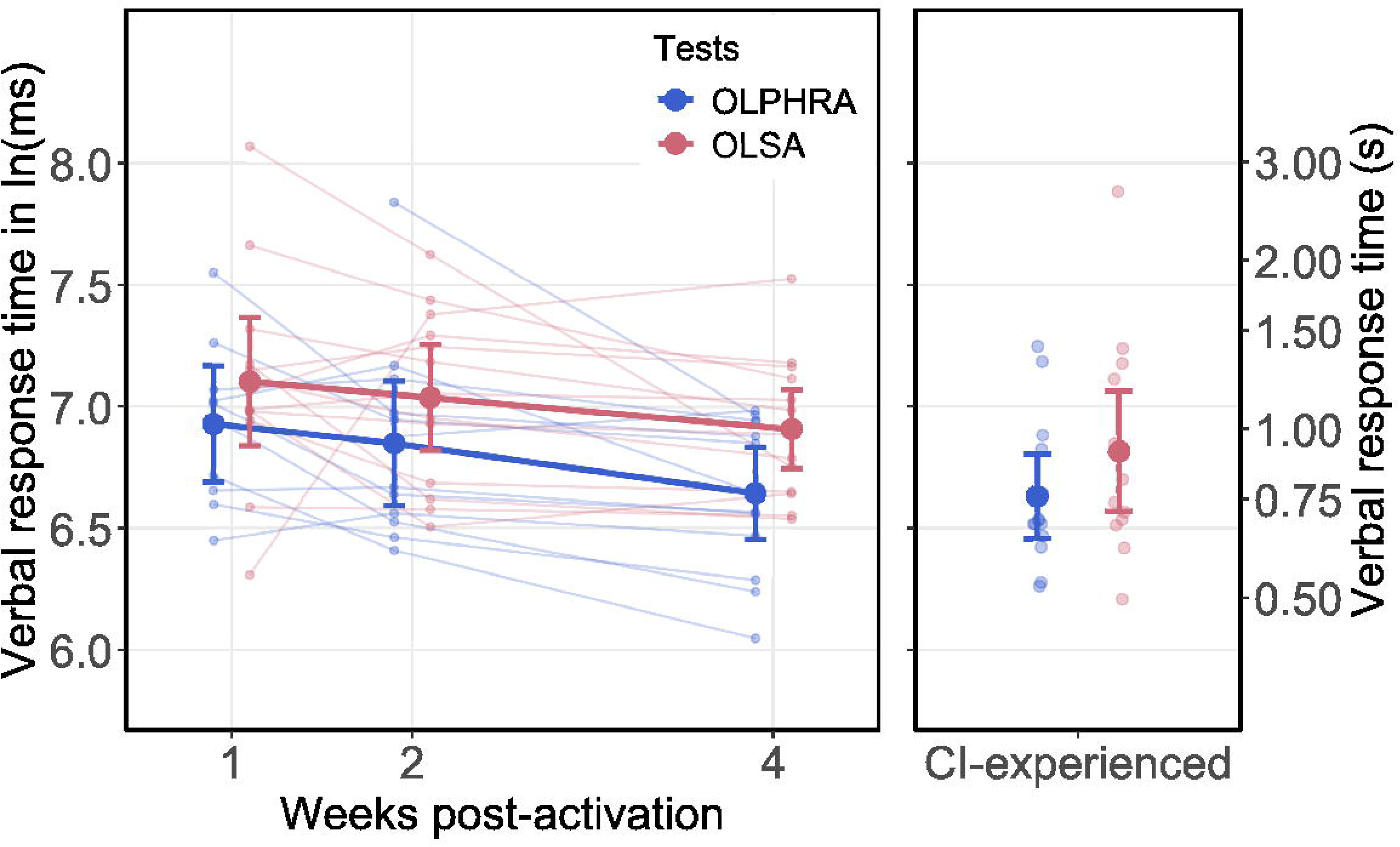
Verbal response times (VRTs) of the CI-rehab group at 1, 2 and 4 weeks after initial fitting (left) and of the CI-experienced group (right) for the OLPHRA and the OLSA. Thin lines and small points show individual participants (valid VRTs only); thick lines and large symbols show the group mean of ln(VRT in ms) with 95% confidence intervals. The right y-axis shows the corresponding response time in seconds.

To compare VRTs between groups, the four-week CI-rehab data were analysed together with the CI-experienced group (55 observations from 28 participants; see Table 5 and 6). VRTs were shorter for the OLPHRA than for the OLSA [*F*(1, 24.4) = 12.32, *p* = .002]. The OLPHRA VRTs were on average 19% shorter than the OLSA VRTs (ratio = 0.81, 95% confidence interval [0.72, 0.92]). VRTs did not differ between CI-rehab and CI-experienced users [*F*(1, 25.0) = 0.34, *p* = .568], and the group-test interaction was not significant [*F*(1, 24.4) = 0.19, *p* = .669]. The advantage of the OLPHRA was thus comparable in both groups. In the CI-rehab group, subjective listening effort was not significantly correlated with VRT at any time point or for either test (Spearman’s ρ = 0.06–0.64, all Benjamini-Hochberg-adjusted p ≥ .11; n = 10–14). In the CI-experienced group, listening effort correlated positively with VRT for the OLSA (ρ = 0.61, n = 14, adjusted *p* = .044) but not for the OLPHRA (ρ = 0.46, adjusted *p* = .099).

**Table 6:**
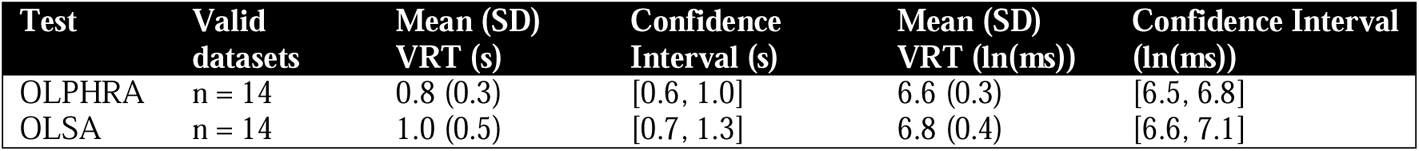
VRTs of the CI-experienced group for each test (OLPHRA, OLSA). VRTs in s and after logarithmic transformation in ln(ms) are given as mean (SD) and 95% confidence interval across valid datasets.

### Pupillometry

To complement speech recognition and VRT, we analysed task-evoked pupil dilation as a physiological index of listening effort. Because of the data loss described in the Methods section, the analysis was restricted to the four-week time point of the CI-rehab group (*n* = 13) and the CI-experienced group (*n* = 10), yielding 46 pupil traces (two tests per participant).

Mean pupil traces did not fully return to baseline before the next trial (Fig 4), reflecting the absence of a fixed silent interval between trials. Thus, results should be regarded as exploratory. Mean pupil dilation and slope within the stimulus window are given per group and test in Tables 7 and 8.

**Fig 4:**
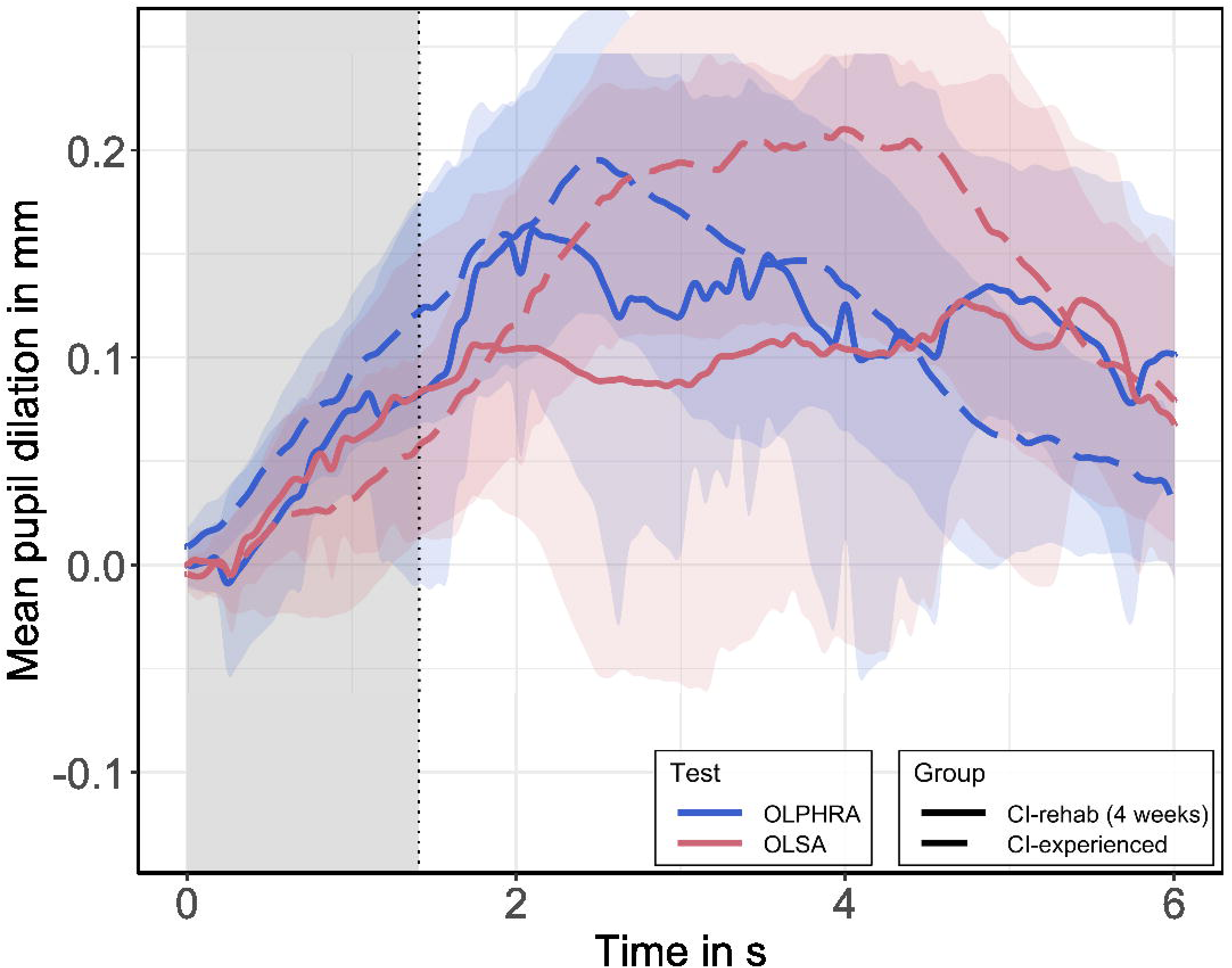
Mean pupil dilation time series (in mm) after baseline correction for the OLPHRA and OLSA in the CI-rehab group at the four-week time point (solid lines, *n* = 13) and the CI-experienced group (dashed lines, *n* = 10). Shaded areas denote the 95% confidence interval of the mean across participants. The grey area marks the stimulus window up to the offset of the shortest stimulus (1.41 s; vertical dotted line). The white area represents the remaining stimulus presentation and response window (up to 6 s).

**Table 7:** Pupillary metrics (mean pupil dilation and slope) within the stimulus window [0 1.41s] of the CI-rehab group (four-week time point) for each test (OLPHRA, OLSA). Mean pupil dilation (in mm) and slopes (in mm/s) as mean (SD) and 95% confidence interval across participants (*n* = 13). Values are descriptive statistics of per-participant linear fits and may differ slightly from the model-based estimates reported in the text.

| **Metric** | **Test** | **Mean (SD)** | **Confidence Interval** |
|---|---|---|---|
| Mean pupil dilation (in mm) | OLPHRA | 0.040 (0.069) | [-0.002, 0.081] |
| Slope (in mm/s) | OLPHRA | 0.078 (0.070) | [0.035, 0.120] |
| Mean pupil dilation (in mm) | OLSA | 0.039 (0.061) | [0.003, 0.076] |
| Slope (in mm/s) | OLSA | 0.068 (0.081) | [0.019, 0.117] |

**Table 8:** Pupillary metrics (mean pupil dilation and slope) within the stimulus window [0 1.41s] of the CI-experienced group for each test (OLPHRA, OLSA). Mean pupil dilation (in mm) and slopes (in mm/s) as mean (SD) and 95% confidence interval across participants (*n* = 10). Values are descriptive statistics of per-participant linear fits and may differ slightly from the model-based estimates reported in the text.

| **Metric** | **Test** | **Mean (SD)** | **Confidence Interval** |
|---|---|---|---|
| Mean pupil dilation (in mm) | OLPHRA | 0.065 (0.037) | [0.038, 0.091] |
| Slope (in mm/s) | OLPHRA | 0.085 (0.057) | [0.044, 0.126] |
| Mean pupil dilation (in mm) | OLSA | 0.023 (0.053) | [-0.015, 0.061] |
| Slope (in mm/s) | OLSA | 0.044 (0.048) | [0.010, 0.079] |

In both groups, pupil dilation rose faster during the OLPHRA than during the OLSA (Fig 4). The GCA confirmed a significant increase in pupil size across the stimulus window [*F*(1, 19.8) = 25.4, *p* < .001; β = 0.069 mm/s] and a significant time × test interaction [*F*(1, 20.3) = 5.66, *p* = .027]. Averaged across groups, the slope was steeper for the OLPHRA (0.080 mm/s) than for the OLSA (0.057 mm/s), a difference of 0.024 mm/s, 95% confidence interval [0.003, 0.045]. No effects involving group were significant (group: *p* = .35; time × group: *p* = .94; test × group: *p* = .20; time × test × group: *p* = .13). Age, which was included as a covariate because the groups differed in age, had no significant effect (age: *p* = .50; time × age: *p* = .42). A sensitivity analysis without age yielded the same time × test interaction [*F*(1, 20.4) = 5.67, *p* = .027] and no significant group effects. GCA model estimates are given in S2 Table.

Mean pupil dilation and slope were strongly correlated across the 46 traces (*r* = .90, 95% confidence interval [.83, .95]). Accordingly, a linear mixed model on mean pupil dilation showed a significant effect of test [*F*(1, 21) = 4.62, *p* = .043] and a borderline test × group interaction [*F*(1, 21) = 4.34, *p* = .050], with no effect of group (*p* = .72) or age (*p* = .64). As Tables 7 and 8 show, the difference between OLPHRA and OLSA was 0.042 mm in the CI-experienced group but only 0.001 mm in the CI-rehab group.

## Discussion

The primary objective of this study was to investigate the clinical utility and feasibility of the novel OLPHRA speech test in individuals with CI, tracking both newly implanted recipients during their initial rehabilitation phase and long-term, experienced CI-users. Beyond conventional behavioural speech recognition metrics, we utilized VRTs and, in an exploratory analysis, task-evoked pupillometry to capture continuous speech processing dynamics and index listening effort during active speech perception.

### Clinical Utility of the OLPHRA in CI patients

Speech-recognition scores for the novel OLPHRA and the established OLSA did not differ significantly, neither in the longitudinal CI-rehab cohort (*p* = .47) nor in the comparison with the CI-experienced group (*p* = .69). As the study was not designed to test equivalence, this indicates similar scores but does not show equal performance. This outcome mirrors the findings of [9], who observed no significant differences in speech recognition thresholds when comparing the OLPHRA to synthesized versions of the OLSA and GÖSA in older hearing-impaired populations. While [9] evaluated speech recognition adaptively in noise, the performance in the current study was observed using fixed-level presentations in quiet with a contralateral masking noise. Despite these differing primary metrics, both datasets indicate that the lexical and structural variations between the phrase-based OLPHRA and the sentence-based OLSA do not result in a systematic divergence in raw speech-recognition accuracy.

These performance similarities occurred despite distinct structural and linguistic differences between the materials. The matrix-style OLSA utilizes a 5-word sentence structure (name– verb–numeral–adjective–noun) that is syntactically fixed but semantically unpredictable. In contrast, the OLPHRA consists of short, 4-word phrases with a fixed syntactic structure (article–adjective–noun–infinitive) providing everyday semantic context. This closer semantic coupling is reflected by the OLPHRA’s lower *j*-factor (*j* = 3.1 at -6 dB SNR) relative to the OLSA sentences (*j* = 4.29 at -5 dB SNR; [9,11]), although these values were determined in earlier studies at different SNRs and not in the present sample.

Despite the clearer semantic architecture of the OLPHRA, the lack of a distinct phrase-level performance benefit may reflect long-term familiarity with the OLSA. This explanation is speculative and was not tested here. The OLSA is integrated into German clinical routines; the CI-experienced cohort undergoes repeated OLSA testing during annual evaluations, and many early-stage candidates encounter it during pre-implantation diagnostics. Conversely, the OLPHRA represented entirely novel speech material to all participants, which may have reduced the expected semantic advantage.

The similar scores are unlikely to result from differences in procedural learning during the session. While the OLSA exhibits a known task-specific training effect of approximately 1 to 2 dB SNR [11] or up to 3 dB SNR [45] within the first two test lists, the OLPHRA demonstrates a negligible training effect of approximately 1 dB SNR across initial measurements, despite being completely unfamiliar to the listeners [9]. Because all participants completed two training lists prior to data collection for both tests, any immediate procedural training effects are unlikely to explain the similar scores. Long-term familiarity with the OLSA therefore remains a possible explanation [46], but this cannot be concluded from the present data.

In contrast to the matrix tests, scores for the open-set FMST were significantly poorer across both cohorts, reflecting several known factors. First, as a single-word test, the FMST lacks linguistic or semantic context to support top-down repairs [6]. Second, evaluations indicate that the underlying test lists do not meet contemporary standards for structural, perceptual, and phonemic equivalence [47]. Additionally, linguistic reviews show that approximately 45% of the original 1953 FMST test words have fallen out of contemporary German usage [48]. Combined with uncalibrated presentation levels across historical recording variants [31], these factors reduce the reliability of the FMST as a metric of modern speech perception. Moreover, the FMST also differed from the matrix tests in procedure (one list, no training lists, no contralateral masker, different booth and loudspeaker distance, natural male voice). The lower FMST scores therefore cannot be attributed to the test material alone and do not demonstrate better measurement properties of the OLPHRA. Despite these limitations, the FMST continues to be routinely administered in clinical settings. Therefore, comparing its results with those of the OLPHRA and OLSA is of considerable clinical relevance.

In both groups, the OLPHRA yielded plausible scores that were similar to those of the OLSA. In older adults with hearing loss, [9] reported a steep psychometric function for the OLPHRA, especially in comparison to the FMST, and the test uses speech tokens that resemble everyday language structures [8]. Furthermore, scores increased from week one to week two and from week two to week four, as they did for the OLSA. This shows that the OLPHRA reflects improvements during early rehabilitation, but not that it is more sensitive than the OLSA. The improvement cannot be separated from processor adjustments, growing familiarity with the test situation, and repeated testing, and the test-retest reliability of the OLPHRA was not assessed. Possible advantages of the OLPHRA over the OLSA are its shorter duration, its everyday material, and the shorter VRTs described below.

However, at the four-week time point, speech-recognition scores did not differ significantly from, but tended to be lower than, those of the CI-experienced cohort (*p* = .09). Ceiling effects were mainly visible in the CI-experienced group, whereas scores in the CI-rehab group were highly variable. The groups also differed in age and device configuration, so this comparison should be interpreted with caution. The maximum accuracy observed in the CI-experienced group suggests that testing in quiet, even with a continuous contralateral masking noise at 75 dB SPL, lacks the difficulty required to track later progress. A ceiling effect indicates limited sensitivity at high-performance levels and does not imply that abilities have converged. Studies with longer follow-up report continued improvement beyond the first weeks of CI use [49], so our conclusions are restricted to the first four weeks in this small sample. To preserve the clinical utility of the test as patient performance and improvement evolve, future protocols could address these ceiling effects by introducing an ipsilateral masking noise (such as test-specific OLnoise or OLPHRAnoise) as recommended in [5] or by transitioning to an adaptive threshold-seeking procedure beyond the initial weeks of device activation.

### Behavioural dynamics of listening effort: VRT

While speech-recognition scores did not differ between the test materials, verbal response times revealed differences in processing speed. Overall, VRTs were shorter for the OLPHRA than for the OLSA (ratio 0.83), with the same direction at all time points. After correction for multiple comparisons, the difference was significant at week four only, and it was also present in the week-four comparison with the CI-experienced group. VRTs also decreased with device experience, but the OLPHRA advantage did not change significantly over time (no time × test interaction).

This difference may be partially driven by structural variations. The OLPHRA consists of four words, whereas the OLSA contains five words, resulting in a mean phrase duration that is approximately 200 ms shorter for the OLPHRA (see Table 2). Fewer lexical units can reduce working memory demands during stimulus retention and recall, facilitating faster response execution upon stimulus offset [18,20]. This interpretation aligns with [21], who recorded VRT and pupil dilation simultaneously during an Italian matrix sentence test and observed that more difficult listening conditions (lower SNRs) systematically prolonged VRTs. In this context, the shorter duration and the predictive semantic structure of the OLPHRA may have contributed to reduced processing difficulty, resulting in faster response execution compared to the semantically unpredictable matrix sentences of the OLSA. This is consistent with the lower *j*-factor of the OLPHRA [8]. However, word count, duration, speaker voice, and semantic context differ between the tests, so the VRT difference cannot be attributed to semantic context alone.

Acoustic properties may also influence these behavioural temporal patterns. The OLPHRA utilizes a synthesized female voice, whereas the OLSA features a natural male speaker. Speaker gender categorization can be difficult for CI users due to altered fundamental frequency (f0) and vocal tract length cues [50]. Five participants spontaneously reported a preference for the female voice; this observation was not recorded systematically.

Perceptually, subjective ratings indicated that participants did not consistently distinguish the synthetic nature of the speech tokens in the OLPHRA. In the CI-rehab group, only about half of the participants classified each test correctly (8/15 for both), which is close to chance level. In contrast, the CI-experienced group showed a clearer differentiation between the two, with OLPHRA more often perceived as synthetic and the OLSA more consistently identified as natural male voice. Although [51] reported poorer recognition for synthesized speech in CI users, we found no indication of a handicap for the synthetic OLPHRA in speech-recognition scores or VRTs. Voice and test material cannot be separated in this design.

Subjective listening effort was positively associated with VRT, but the association was mostly not significant. After correction for multiple comparisons, only the OLSA in the CI-experienced group showed a significant correlation (ρ = .61, adjusted *p* = .044). All other correlations were not significant, although all eight had a positive sign (ρ = .06–.64). This pattern agrees with [19], who observed that longer response latencies went along with higher self-reported effort, and only partly with [18], who found no close link between the two measures. Given the small samples (n = 10–14 per test), an adjusted p value close to .05, and a correction family of only two tests in the CI-experienced group, the single significant result is exploratory and needs replication. Differences in data processing may also contribute to the divergent findings in the literature. [19] z-transformed the log-transformed VRTs and normalised the effort ranks within each participant across several conditions. This removes individual differences in baseline response speed and in the use of the rating scale. Because each measurement interval of the present study comprised only two test conditions, such a within-participant normalisation was not feasible, and between-participant baseline differences remained in the data. These differences may have attenuated the correlations. Whether VRT directly indexes listening effort or partly reflects other processes, such as lexical retrieval confidence and response hesitation, therefore remains open.

### Pupillary Dynamics and Temporal Trajectories of Speech Processing

These pupillometric results are exploratory because of the small sample size (13 CI-rehab participants at week four and 10 CI-experienced participants). In addition to the VRT metric, pupillary dynamics provided a physiological index of listening effort. Utilizing a mixed-effects GCA including both groups with group as a factor, we show that the linear trajectory of pupil dilation was steeper for the OLPHRA than for the OLSA (difference 0.024 mm/s, 95% confidence interval [0.003, 0.045]). No effects involving group were significant, but the difference was visibly larger in the CI-experienced group (Fig 4, Tables 7 and 8), and the test × group interaction for mean pupil dilation was borderline (*p* = .050). We therefore interpret group differences with caution. Previous studies have suggested that the initial slope leading to maximum pupil expansion represents the speed and intensity of immediate auditory resource allocation during signal encoding [27,28]. Furthermore, [21] demonstrated that alterations in cognitive demands during speech recognition systematically alter pupillary time courses. Applying this framework to our GCA findings, the steeper slope for the OLPHRA may reflect a faster recruitment of cognitive resources during active signal encoding.

However, mean pupil dilation and slope were strongly correlated (*r* = .90) and mean dilation was also higher for the OLPHRA, so a higher processing demand cannot be excluded. The result contradicts our hypothesis (a) of a flatter slope for the OLPHRA. Because the OLPHRA is shorter and more predictable in syntactic structure, it may facilitate more efficient allocation of cognitive resources during speech comprehension, but this remains speculative.

The pupil slope and the VRT results point in different directions relative to our hypothesis: the OLPHRA elicited shorter VRTs but a steeper pupil slope. Whether the two measures capture different aspects of listening effort, as suggested by [21], cannot be concluded from our data, because their relation was not analysed. Finally, the present results show that continuous temporal profiles of the pupillary responses diverge between the two tests. For the OLPHRA, the pupil response peaks near the mean duration of the phrases, a timeline consistent with conventional pupillometric patterns in speech tasks [25,29]. In contrast, the OLSA response appears to climb steadily to a peak at approximately 3-4 s post-stimulus onset, extending considerably longer into the response window. These peak latencies are visual observations from Fig 4 outside the analysed window and were not tested statistically. While a later peak is expected for longer stimuli, the longest OLSA sentence ends at 2.77 s, leaving a substantial delay before the maximum pupillary response is reached. Peak timing is also confounded with speech production and response timing, which differ between tests, and the pupil did not return to baseline between trials.

This temporal shift may indicate different processing and recall strategies between the two tasks, extending the findings of [29]. They compared peak pupil dilations across different speech tests in CI users and demonstrated that structural variations between meaningful, everyday sentence material and semantically unpredictable matrix sentences alter physiological effort profiles. In the current study, this architectural difference may affect the temporal resolution of resource allocation. In the matrix-style OLSA, listeners must hold and match incoming acoustic tokens against a constrained, closed-set vocabulary repertoire, which could delay definitive integration until the full matrix is received. In the OLPHRA, listeners process an open-set, but semantically coherent and predictive phrase. Although both tests utilize structured layouts, decoding the highly predictable phrases of the OLPHRA may allow for rapid, immediate semantic reconstruction during stimulus presentation, explaining the highly accelerated initial GCA trajectory observed during active OLPHRA perception.

### Methodological Considerations and Limitations

Several methodological factors and limitations should be considered when interpreting these findings. First, testing occurred immediately following regular clinical appointments, which differed in duration and cognitive demand. The CI-rehab group completed a 3-hour rehabilitation session comprising audiometric testing, hearing therapy, and device adjustments, while the CI-experienced group underwent a standard 2-hour annual assessment, including audiometric testing and CI fitting. Consequently, baseline fatigue levels prior to the experiment were likely not comparable between cohorts. The groups also differed in age (the CI-rehab group was older) and in device configuration. Comparisons between groups were cross-sectional and based on small samples, so they do not allow conclusions about equivalence or about the development of individual users. No sample size calculation was performed, and the study was not powered for the group comparisons. Participants were recruited at a single centre, with heterogeneous device configurations; results may not generalise to other CI populations or languages.

For newly implanted participants, the testing environment also represented an unfamiliar auditory and technical situation. Many individuals in the CI-rehab group had experienced prolonged sensory deprivation in the implanted ear prior to surgery, making initial acclimatisation to electrical stimulation a cognitive challenge. Furthermore, during these initial weeks, programming adjustments to the speech processor map are larger and more frequent than the stable settings of long-term users. Managing these changing acoustic boundaries while the non-implanted ear is potentially impaired can introduce fatigue, which may have influenced the longitudinal behavioural data.

A second limitation relates to the heterogeneous device configurations across both cohorts, which included bilateral, bimodal, and SSD fittings (CI-rehab: 11 bimodal, 3 bilateral, 1 SSD; CI-experienced: 3, 8, and 3). Although continuous contralateral masking noise was applied at 75 dB SPL to control the non-test ear, the effective perception of this noise likely varied. We did not verify effective masking, and residual hearing and the device status of the non-test ear were not controlled beyond the protocol. Participants with SSD may have perceived the masking noise as louder or more disruptive than bilaterally or bimodally fitted individuals with residual hearing. Such differences in masking perception can influence cognitive load and speech-recognition performance, as higher noise levels are associated with lower speech-recognition scores, prolonged verbal response times [19,20] and increased pupil dilation profiles [19,20,24,29,30]. The masker may thus also have affected the pupil responses directly. In addition, test material, speaker voice, word count, duration, and familiarity differed between OLPHRA and OLSA, and training in the best-aided condition differed from the measurement condition.

Third, the simultaneous recording of VRTs and pupillometric responses required a trial design without a post-stimulus retention interval [21]. This constraint meant that processing load could only be isolated during the active presentation phase, rather than during a distinct retention or recall window. Because the sentences and phrases varied in duration, with the OLSA sentences being longer than the OLPHRA phrases (see Table 2), defining the pupillometric analysis window presented a challenge. To ensure data consistency across all conditions, a window matching the shortest overall stimulus duration (1.41 s) was selected, restricting the formal GCA to the initial encoding slope. Because of data loss, only 23 of 29 participants could be analysed and only 6 CI-rehab participants had valid pupil data at all three time points, so no longitudinal pupil analysis was possible.

Consequently, peak pupil dilation could not be evaluated because the maximum pupillary response consistently shifted into the active listening and response window. Furthermore, differences in mean pupil dilation within this restricted window were small and were mainly present in the CI-experienced group. To approximate standard clinical routines, a fixed response interval was not enforced between trials; instead, the next stimulus was initiated 2 to 3 s after the participant’s vocalisation ceased. In several instances, this interval was insufficient to allow the pupil trace to fully trend back to a resting state which may have contributed to elevated baseline pupil sizes. A baseline located on a falling flank of the preceding response, or carry-over between trials, may also have influenced the early slope.

These methodological challenges raise a practical question for future clinical protocols regarding whether both metrics are required simultaneously. The current data do not allow a conclusion on whether VRT and pupil slope reflect the same or different aspects of listening effort. However, implementing a pupillometric setup in standard clinical care demands specialised equipment, controlled lighting environments, and lengthy calibration procedures. In contrast, tracking VRTs requires only a microphone setup and minimal timing software. Given its logistical simplicity and sensitivity to test material differences, VRT is easier to implement and may merit further evaluation as a supplementary measure alongside standard speech-recognition scores. Its validity and reliability for clinical use remain to be established.

## Conclusion

This study examined the feasibility of the novel OLPHRA in newly fitted and experienced CI users, using speech-recognition scores, verbal response times (VRTs) and, in an exploratory analysis, task-evoked pupil dilation. Consistent with hypothesis (b), speech-recognition scores increased over the first four weeks of device use for all three tests. OLPHRA scores did not differ significantly from OLSA scores, and at week four they did not differ significantly from those of the CI-experienced group, although the CI-rehab group tended to score lower. As the study was not designed to test equivalence, these results indicate similar scores but not equal performance. They suggest that the OLPHRA can be administered in early and long-term CI users and yields scores similar to the OLSA. They do not show greater sensitivity than the OLSA, and its reliability and clinical validity remain to be established. Scores of the CI-experienced group were close to ceiling, so for later follow-up the test should be combined with ipsilateral masking noise or an adaptive procedure. The improvement over four weeks cannot be separated from processor adjustments, growing familiarity with the setting and repeated testing, so conclusions are restricted to the first four weeks in this small sample.

Hypothesis (a) was only partly supported. OLPHRA scores were not higher than OLSA scores, but VRTs were shorter for the OLPHRA overall (ratio 0.83, 95% confidence interval [0.74, 0.93]), with the same direction at all time points and a significant difference at week four. VRTs also decreased with device experience, as expected in hypothesis (c), whereas the OLPHRA advantage did not change significantly over time. Because word count, duration, speaker voice and semantic context differ between the tests, the shorter VRTs cannot be attributed to semantic context alone. In the exploratory pupillometric analysis, the initial pupil slope was steeper for the OLPHRA than for the OLSA, contrary to hypothesis (a). This may reflect faster recruitment of cognitive resources, but a higher processing demand cannot be excluded. Given the small sample, the short analysis window and the incomplete return to baseline between trials, this finding is hypothesis-generating. Pupil slopes could not be analysed longitudinally, so this part of hypothesis (c) remains untested. The relation between VRT and pupil measures was not analysed, so no conclusion can be drawn on whether they capture different aspects of listening effort. VRT is easy to record and may merit further evaluation as a supplementary measure alongside speech-recognition scores. Future studies should use larger samples, adaptive or ipsilateral-noise conditions, comparable stimulus durations and a controlled inter-trial interval.

## Supporting information

S4 Checklist

## Data Availability

All data produced are available online at https://doi.org/10.5281/zenodo.23727483

## Acknowledgements

The authors acknowledge the Hörzentrum Oldenburg gGmbH, Oldenburg, Germany for providing the speech test recordings of the Oldenburg Sentence Test.

## Supporting information

S1 Table. Linear mixed models of log-transformed verbal response times.

S2 Table. Growth curve models of pupil dilation

S3 Table. Linear mixed model of mean pupil dilation.

S4 Checklist. STROBE checklist for observational studies.

## Notes

### Competing Interest Statement

The authors have declared no competing interest.

### Clinical Trial

German Clinical Trials Register DRKS00040892

### Author Declarations

Ethics Committee of Carl von Ossietzky Universitaet Oldenburg gave ethical approval for this work (approval number: 2021-095)

