## Supplementary material for "Feasibility of the Oldenburg phrase test (OLPHRA) in cochlear-implant users": S1 Table

**S1 Table:** Linear mixed models of natural-log-transformed verbal response times (VRT; ln of VRT in ms). Both models included a random intercept per participant; fixed effects were time point, test (OLPHRA, OLSA) and their interaction (A, B) or group, test and their interaction (C, D). Degrees of freedom and p values of the F tests according to Satterthwaite. Ratios are back-transformed contrasts of estimated marginal means on the log scale (OLPHRA / OLSA, or the later vs. the earlier time point as indicated), obtained with emmeans; values below 1 indicate shorter VRTs for the numerator. The p values of the post-hoc contrasts are adjusted with the Benjamini-Hochberg procedure, the 95% confidence intervals (CI) are unadjusted. The overall OLPHRA / OLSA contrasts (A, D) are single comparisons, not adjusted, and tested by the F test of test above.

| **Effect / contrast** | **F** | **df** | **Ratio** | **95% CI** | **p** |
| --- | --- | --- | --- | --- | --- |
| **A. CI-rehab group, four-week time course (75 observations from 14 participants): F tests** | | | | | |
| Time point | 8.14 | 2, 57.1 |  |  | < .001 |
| Test | 10.30 | 1, 57.3 |  |  | .002 |
| Time point × test | 0.60 | 2, 56.5 |  |  | .55 |
| **B. CI-rehab group: ratios of geometric mean VRTs** | | | | | |
| OLPHRA / OLSA, averaged across time points |  |  | 0.83 | [0.74, 0.93] | – |
| OLPHRA / OLSA, week 1 |  |  | 0.90 | [0.73, 1.11] | .303 |
| OLPHRA / OLSA, week 2 |  |  | 0.83 | [0.68, 1.01] | .096 |
| OLPHRA / OLSA, week 4 |  |  | 0.77 | [0.64, 0.93] | .023 |
| Week 1 / week 4 |  |  | 1.32 | [1.15, 1.52] | < .001 |
| Week 2 / week 4 |  |  | 1.19 | [1.04, 1.37] | .020 |
| Week 1 / week 2 |  |  | 1.11 | [0.96, 1.28] | .149 |
| **C. CI-rehab (week 4) vs. CI-experienced (55 observations from 28 participants): F tests** | | | | | |
| Group | 0.34 | 1, 25.0 |  |  | .568 |
| Test | 12.32 | 1, 24.4 |  |  | .002 |
| Group × test | 0.19 | 1, 24.4 |  |  | .669 |
| **D. Ratio of geometric mean VRTs** | | | | | |
| OLPHRA / OLSA, averaged across groups |  |  | 0.81 | [0.72, 0.92] | – |
