## Supplementary material for "Feasibility of the Oldenburg phrase test (OLPHRA) in cochlear-implant users": S2 Table

**S2 Table:** Growth curve models of pupil dilation within the stimulus window (0–1.41 s; 100-ms bins; 690 observations from 46 participant × test combinations of 23 participants). The primary model included age as a covariate; the sensitivity model did not. Time in s, pupil size in mm, age grand-mean centred. Test and group were sum-coded (OLPHRA = +1, OLSA = −1; CI-rehab = +1, CI-experienced = −1). Degrees of freedom according to Satterthwaite (primary model: 19.8–41.0; sensitivity model: 20.4–42.0). The 95% confidence intervals (CI) of the fixed effects are t-based, calculated from the estimates, standard errors and Satterthwaite degrees of freedom; degrees of freedom for model-based slopes according to Kenward-Roger.

| **Term** | **Primary model: Estimate [95% CI]** | **t** | **p** | **Sensitivity model: Estimate [95% CI]** | **t** | **p** |
| --- | --- | --- | --- | --- | --- | --- |
| **A. Fixed effects** | | | | | | |
| Intercept | −0.0065 [−0.0140, 0.0010] | −1.75 | .088 | −0.0064 [−0.0138, 0.0011] | −1.73 | .091 |
| Time | 0.0685 [0.0401, 0.0969] | 5.04 | < .001 | 0.0679 [0.0399, 0.0960] | 5.04 | < .001 |
| Test | 0.0021 [−0.0054, 0.0096] | 0.57 | .574 | 0.0021 [−0.0053, 0.0095] | 0.57 | .572 |
| Group | −0.0038 [−0.0118, 0.0043] | −0.94 | .352 | −0.0047 [−0.0122, 0.0027] | −1.29 | .204 |
| Age | −0.0002 [−0.0007, 0.0003] | −0.67 | .505 | – | – | – |
| Time × test | 0.0119 [0.0015, 0.0223] | 2.38 | .027 | 0.0119 [0.0015, 0.0223] | 2.38 | .027 |
| Time × group | −0.0011 [−0.0316, 0.0294] | −0.08 | .941 | 0.0033 [−0.0248, 0.0314] | 0.25 | .809 |
| Test × group | −0.0048 [−0.0123, 0.0027] | −1.29 | .203 | −0.0048 [−0.0122, 0.0026] | −1.30 | .200 |
| Time × age | 0.0008 [−0.0012, 0.0027] | 0.82 | .424 | – | – | – |
| Time × test × group | −0.0079 [−0.0183, 0.0025] | −1.58 | .129 | −0.0079 [−0.0183, 0.0025] | −1.58 | .129 |
| **B. Random effects (variance)** | | | | | | |
| Participant × test: intercept | 0.00047 |  |  | 0.00046 |  |  |
| Participant × test: time slope | 0.00093 (r = .31) |  |  | 0.00093 (r = .30) |  |  |
| Participant: time slope | 0.0036 |  |  | 0.0035 |  |  |
| Residual | 0.00055 |  |  | 0.00055 |  |  |
| **C. Model-based slopes in mm/s [95% confidence interval]** | | | | | | |
| CI-rehab, OLPHRA | 0.071 [0.031, 0.112] |  |  | 0.075 [0.036, 0.114] |  |  |
| CI-rehab, OLSA | 0.063 [0.023, 0.104] |  |  | 0.067 [0.028, 0.106] |  |  |
| CI-experienced, OLPHRA | 0.089 [0.043, 0.136] |  |  | 0.085 [0.040, 0.129] |  |  |
| CI-experienced, OLSA | 0.050 [0.003, 0.096] |  |  | 0.045 [0.000, 0.089] |  |  |
