## Supplementary material for "Feasibility of the Oldenburg phrase test (OLPHRA) in cochlear-implant users": S3 Table

**S3 Table:** Linear mixed model of mean pupil dilation within the stimulus window (0–1.41 s) in the four-week CI-rehab group (n = 13) and the CI-experienced group (n = 10; 46 traces). Fixed effects were test (OLPHRA, OLSA), group (CI-rehab, CI-experienced), their interaction and age (grand-mean centred), with a random intercept per participant. Test and group were sum-coded. Degrees of freedom and p values according to Satterthwaite. This analysis was exploratory. Estimated means were obtained with emmeans.

| **Effect** | **F** | **df** | **p** |
| --- | --- | --- | --- |
| **A. Fixed effects (F tests)** | | | |
| Test | 4.62 | 1, 21 | .043 |
| Group | 0.13 | 1, 20 | .72 |
| Test × group | 4.34 | 1, 21 | .050 |
| Age | 0.23 | 1, 20 | .64 |
| **B. Estimated mean pupil dilation in mm [95% confidence interval]** | | | |
| CI-rehab, OLPHRA | 0.038 [0.004, 0.072] |  |  |
| CI-rehab, OLSA | 0.037 [0.003, 0.072] |  |  |
| CI-experienced, OLPHRA | 0.067 [0.028, 0.107] |  |  |
| CI-experienced, OLSA | 0.025 [−0.014, 0.065] |  |  |
| **C. Association of mean pupil dilation and slope (46 traces)** | | | |
| Pearson correlation r [95% confidence interval] | .90 [.83, .95] |  |  |
