## Supplementary material for "Feasibility of the Oldenburg phrase test (OLPHRA) in cochlear-implant users": S4 Checklist

STROBE Checklist

| Item | Present | Page |
| --- | --- | --- |
| 1.a Title and abstract | ☒ | 1 |
| 1.b Summary and objective information on what was done and found in the abstract | ☒ | 1 |
| 2.a Scientific background of the study | ☒ | 2 |
| 2.b Rationale of the study | ☒ | 4 |
| 3.a Specific objectives of the study | ☒ | 4 |
| 3.b A priori hypotheses of the study | ☒ | 5 |
| 3.c Research question | ☒ | 4 |
| 4. Study design | ☒ | 5 |
| 5. Setting, locations, relevant dates, periods of recruitment, exposure, follow-up and data collection | ☒ | 6 |
| 6. Description of the population adapted to the type of study | ☒ | 5 |
| 7. Variables: clear description of outcome criteria, exposure, prediction factors, potential confounding factors, etc. | ☒ | 9 |
| 8. Indication of data sources and evaluation methods for each variable of interest | ☒ | 9 |
| 9. Description of all measures taken to avoid potential sources of bias | ☒ | 10 |
| 10. Explanation of the number of subjects to include | ☒ | 5 |
| 11. Treatment of quantitative variables and justification | ☒ | 11 |
| 12.a Description of all statistical analyses | ☒ | 13 |
| 12.b Description of methods used | ☒ | 10ff |
| 12.c Description of missing data treatment | ☒ | 10ff |
| 12.d Description of specific data treatment by study type (lost to follow-up, case-control matching, sampling) | ☒ | 10 |
| 12.e Description of sensitivity analyses | ☒ | 15 |
| 13. Description of the population at each stage of the study, number of individuals, reasons for non-participation, presence of a flow diagram | ☒ | 5 |
| 14.a Description of the characteristics of the studied population | ☒ | 5 |
| 14.b Description of the number of subjects and missing data for each variable | ☒ | 10-12 |
| 14.c For cohort studies, summary of the follow-up period | ☒ | 9 |
| 15. Description of obtained data (adapted to study type) (e.g., number of events, number of exposed or unexposed subjects) | ☒ | 9-10 |
| 16.a Description of main results: unadjusted, adjusted estimates, presence of confidence interval | ☒ | 16ff |
| 17. Description of other analyses performed: subgroup analysis, sensitivity analysis | ☒ | 20-22 |
| 18. Summary of main results | ☒ | 32-33 |
| 19. Discussion of study limitations | ☒ | 29 |
| 20. General interpretation of results taking into account objectives, study limitations, etc. | ☒ | 22-31 |
| 21. Discussion of the generalizability of study results | ☒ | 30 |
| 22. Source of funding | ☒ |  |
